# Evaluation of the Clinical Utility of DxGPT, a GPT-4 Based Large Language Model, through an Analysis of Diagnostic Accuracy and User Experience

**DOI:** 10.1101/2024.07.23.24310847

**Authors:** Marina Alvarez-Estape, Ivan Cano, Rosa Pino, Carla González Grado, Andrea Aldemira-Liz, Javier Gonzálvez-Ortuño, Juanjo do Olmo, Javier Logroño, Marcelo Martínez, Carlos Mascías, Julián Isla, Jordi Martínez Roldán, Cristian Launes, Francesc Garcia-Cuyas, Paula Esteller-Cucala

**Affiliations:** Data and Digital Strategy Department, Hospital Sant Joan de Déu, Barcelona, Spain; Metabolomics and Biochemical Genetics, Institut de Recerca Sant Joan de Déu, Barcelona, Spain; Pediatrics Department, Hospital Sant Joan de Déu, Barcelona, Spain; Environment Effects on Child/Adolescent Well-being, Institut de Recerca Sant Joan de Déu, Barcelona, Spain; Departament de Cirurgia i Especialitats Medicoquirúrgiques, Facultat de Medicina i Ciències de la Salut, Universitat de Barcelona, Barcelona, Spain; Strategy and Planning Department, Hospital Sant Joan de Déu, Barcelona, Spain; Foundation 29, Madrid, Spain; Microsoft, Industry Solution Delivery, Madrid, Spain; Pediatric Infectious Diseases and Microbiome Research Group, Institut de Recerca Sant Joan de Déu, Spain; CIBER Epidemiology and Public Health (CIBERESP), Madrid, Spain

## Abstract

**Importance:** The time to accurately diagnose rare pediatric diseases often spans years. Assessing the diagnostic accuracy of an LLM-based tool on real pediatric cases can help reduce this time, providing quicker diagnoses for patients and their families.

**Objective:** To evaluate the clinical utility of DxGPT as a support tool for differential diagnosis of both common and rare diseases.

**Design:** Unicentric descriptive cross-sectional exploratory study. Anonymized data from 50 pediatric patients’ medical histories, covering common and rare pathologies, were used to generate clinical case notes. Each clinical case included essential data, with some expanded by complementary tests.

**Setting:** This study was conducted at a reference pediatric hospital, Sant Joan de Déu Barcelona Children’s Hospital.

**Participants:** A total of 50 clinical cases were diagnosed by 78 volunteer doctors (medical diagnostic team) with varying experience, each reviewing 3 clinical cases.

**Interventions:** Each clinician listed up to five diagnoses per clinical case note. The same was done on the DxGPT web platform, obtaining the Top-5 diagnostic proposals. To evaluate DxGPT’s variability, each note was queried three times.

**Main Outcome(s) and Measure(s):** The study mainly focused on comparing diagnostic accuracy, defined as the percentage of cases with the correct diagnosis, between the medical diagnostic team and DxGPT. Other evaluation criteria included qualitative assessments. The medical diagnostic team also completed a survey on their user experience with DxGPT.

**Results:** Top-5 diagnostic accuracy was 65% for clinicians and 60% for DxGPT, with no significant differences. Accuracies for common diseases were higher (Clinicians: 79%, DxGPT: 71%) than for rare diseases (Clinicians: 50%, DxGPT: 49%). Accuracy increased similarly in both groups with expanded information, but this increase was only stastitically significant in clinicians (simple 52% vs. expanded 69%; *p*=0.03). DxGPT’s response variability affected less than 5% of clinical case notes. A survey of 48 clinicians rated the DxGPT platform 3.9/5 overall, 4.1/5 for usefulness, and 4.5/5 for usability.

**Conclusions and Relevance:** DxGPT showed diagnostic accuracies similar to medical staff from a pediatric hospital, indicating its potential for supporting differential diagnosis in other settings. Clinicians praised its usability and simplicity. These tools could provide new insights for challenging diagnostic cases.

**Key Points:** *Question:* Is DxGPT, a large language model-based (LLM-based) tool, effective for differential diagnosis support, specifically in the context of a clinical pediatric setting?

*Findings:* In this unicentric cross-sectional study, diagnostic accuracy, measured as the proportion of clinical cases where any of the five diagnostic options included the correct diagnosis, showed comparable results between clinicians and DxGPT. Top-5 accuracy was 65% for clinicians and 60% for DxGPT.

*Meaning:* These findings highlight the potential of LLM-based tools like DxGPT to support clinicians in making accurate and timely diagnoses, ultimately improving patient care.

## Introduction

The accurate diagnosis of pediatric conditions is a cornerstone of effective clinical practice, yet it remains a challenging task, particularly with the complexity of rare diseases. Clinicians often face difficulties in diagnosing both common and rare diseases due to the vast array of possible conditions and the subtlety of symptoms. The delay in diagnosis, which is especially prevalent in pediatric rare diseases and the potential for incorrect diagnosis or non-diagnosis can lead to unnecessary and potentially harmful treatments ^1^.

Artificial intelligence (AI) has emerged as a transformative force across various fields, including healthcare where large language models (LLMs) in particular, hold the potential of improving diagnosis, treatment and patient care ^2,3^. In this regard, specific LLM-based diagnostic tools, like DxGPT ^4^ have been specifically designed to assist in diagnosing a wide spectrum of diseases, from the most common to the rarest. DxGPT (http://dxgpt.app) is an open diagnostic decision support tool powered by GPT-4 and developed by the non-profit organization Foundation 29. By integrating extensive medical knowledge and language processing capabilities of modern LLMs, DxGPT aims to support clinicians in making accurate and timely diagnoses.

Given the promising capabilities of LLMs in healthcare, it is crucial to evaluate these tools against the performance of human clinicians. Such assessments not only validate these models’ effectiveness but also ensure its reliability and safety in clinical settings. Rigorous evaluation is particularly important for tools like DxGPT, which must demonstrate both high diagnostic accuracy and user-friendly integration into clinical workflows ^5^.

This study aims to assess the potential of the DxGPT platform (a tool based on GPT-4) as a support tool for differential diagnosis in real clinical scenarios. For that, we compared the diagnostic accuracy of DxGPT and clinicians using 50 real pediatric clinical cases from a reference children’s hospital. We also evaluated the effect of additional information on diagnostic accuracy and the variability of DxGPT in terms of the consistency of its diagnoses. Finally, we also examined the usability of this tool by clinicians.

## Methods

### Study design

An unicentric descriptive cross-sectional study was conducted in a reference paediatric hospital (Sant Joan de Déu Barcelona Children’s Hospital, University of Barcelona). The data team extracted fully anonymized data belonging to medical histories of 50 patients with confirmed diagnoses. The number of cases selected per medical specialty was proportional to their occurrence at the hospital in 2023 and was gender balanced (eTables 1 from Supplement 2 and eTables 2 and 3 from Supplement 1). This ensured a diverse representation of patients, including gastroenterological, neurological, metabolic, oncological, infectious, nephrological, rheumatological, immunological, endocrinological, cardiological, and dermatological conditions. At least, half of the cases would correspond to a rare disease (with Orphanet’s prevalence ranging from 1:10,000 to 1:100,000).

### Data extraction and case description

To summarize the clinical cases for this study, the data team extracted relevant information from the electronic health record, preferably from the earliest consultation to the hospital, regardless of whether it was an ambulatory consultation or an admission. Each clinical case contained a minimum set of information, including: (i) age and gender, (ii) a description of the main symptoms/signs, (iii) previous illnesses, allergies, surgeries, (iv) data from physical examinations, and (v) results from complementary tests if these were available within the first 72 hours after being requested. Additional data from further complementary tests were added to an expanded version of each clinical case (referred to as extended cases). These complementary tests could include analytical results (from blood, urine, or other body fluid tests) and/or radiological interpretations (derived from radiological images, computed tomography, or magnetic resonance). All clinical cases were presented in Spanish.

A medical research team (MRT), composed of 5 pediatricians with at least 6 years of experience in hospital pediatrics, ensured that the case description included the minimum data necessary for the known diagnosis. They added further data from subsequent consultations when required. Additionally, they assigned a qualitative complexity level (low, medium, or high) to each case by majority, based on the deviation of the actual case from an academic description of a clinical case of the corresponding disease (eTable 4 from Supplement 1).

Out of the 50 clinical cases, 20 also included extended information, thus resulting in 70 clinical cases. Simple and extended clinical cases were evaluated as independent cases unless stated otherwise.

### Participant recruitment and case assessment procedures

A broad invitation was extended to professionals from the hospital’s Pediatrics, Emergency Care, and Intensive Care Unit Departments to voluntarily and anonymously participate in a clinical case marathon as part of the medical diagnostic team (MDT).

The 50 clinical cases were randomized into 50 unique diagnosis forms, each containing 3 cases with varied characteristics such as disease type, medical specialty, complexity, and information level, ensuring diversity in each set.

MDT respondents indicated their professional experience and provided the primary and up to four differential diagnoses for each clinical case. Each respondent diagnosed 3 cases, with extended versions diagnosed twice to ensure comprehensive assessment.

Each clinical case was also queried in DxGPT (which uses GPT-4 0613; DxGPT was accessed between 22-27 March 2024), retrieving the top five diagnoses (Top-5). To evaluate DxGPT’s response variability, each case was queried 3 times, with all responses recorded and evaluated by the MDT.

### User experience evaluation

Additionally, the MDT was asked to voluntarily and anonymously test DxGPT and respond to a questionnaire about their experience (UX questionnaire). This questionnaire included 3 5-point scale questions and three open-ended questions (eMethods from Supplement 1). Answers to the UX questionnaire were analyzed to obtain the distribution and mean score for each of the numerical questions. ChatGPT-3.5 was used to summarize the main findings on the 3 open-ended questions.

### Performance evaluation

All analyses were performed using R version 4.3.1. All code is available at https://github.com/maralest/DxGPT_HSJD_Analysis.

#### Diagnostic accuracy

The diagnostic accuracy for each clinical case was assessed for DxGPT and for the MDT (also referred as clinicians) (Top-5 accuracy). The MRT evaluated whether the final diagnosis was within the Top-5 answers at each diagnostic group. All evaluations made by the MRT were blinded and reached by majority consensus. Each clinical case underwent 2 independent assessments: (i) a dichotomous evaluation, assigning a score of 1 if the correct diagnosis was listed among the provided answers and 0 if it was not, and (ii) a qualitative evaluation using a 4-point Likert scale based on predefined criteria:

∎ 1: incoherent diagnosis list; the suggested diagnoses are not relevant or plausible given the clinical information available
∎ 2: coherent list, with plausible diagnoses, but the necessary diagnostic tests would not yield the precise diagnosis
∎ 3: coherent diagnosis list, not including the precise diagnosis; however, the list suggested a diagnostic approach likely to lead to the precise diagnosis
∎ 4: coherent diagnosis list, including the specific diagnosis (equivalent to a score of 1 in the dichotomous evaluation)

For the dichotomous evaluation, the percentage of correct diagnoses was calculated and compared between the diagnostic groups (DxGPT and clinicians) and disease frequency types (common and rare). Two-sided Clopper-Pearson confidence intervals were calculated, and statistical significance was tested using Fisher’s exact test with Bonferroni correction. Given the differing number of responses between clinicians (eFigure 1 from Supplement 1) and DxGPT (n = 1) for each clinical case, we calculated the mode of the clinicians’ dichotomous scores for each clinical case. This approach ensured an equal number of results between diagnostic groups, thus allowing paired comparisons. Statistical significance between diagnostic groups was tested using McNemar’s Chi-squared Test for all (N=70), rare (N=35) and common clinical cases (N=35).

To study whether the extended information on the 20 cases increased diagnostic accuracy, McNemar’s Chi-squared Test was performed. Additionally, to evaluate differences in the case complexity categories assigned by the MRT, Fisher’s exact Test with Bonferroni correction was used.

We used the Cochran-Armitage Test to examine differences in the proportions of responses on the 4-point Likert scale between diagnostic groups, between the mode of clinicians’ responses and DxGPT and to compare simple against extended cases.

#### DxGPT variability

We evaluated the variability of the Top-5 (set of 5 diagnoses) and Top-1 (first diagnosis) results across multiple responses for the same clinical case. For each clinical case, we calculated the mean of the three-way paired Jaccard coefficient. A Jaccard coefficient of 1 indicates complete overlap between the three sets of Top-5 diagnoses, while a coefficient of 0 indicates no overlap between the 3 replicates.

### Ethical Considerations

The study complies with the Helsinki Declaration and relevant legal requirements. It was approved by the Ethics Committee and Institutional Review Board of the Sant Joan de Déu Hospital (PIC-20-24) and informed consents from patients’ parents were waived. The ethical aspects of the participation by the medical diagnostic team emphasized the complete anonymity of participants, the non-collection of identifiable personal data, and the voluntary nature of their participation.

## Results

### Performance evaluation of clinicians and DxGPT

A total of 78 clinicians, including specialists and residents in pediatrics with varied experience, participated in the case diagnosis (Figure 1 and eFigure 1 from Supplement 1). As each clinician was asked to provide a main diagnosis and up to four alternative diagnoses for three clinical cases, 328 responses were collected (eFigure 1 from Supplement 1).

**Figure 1.**
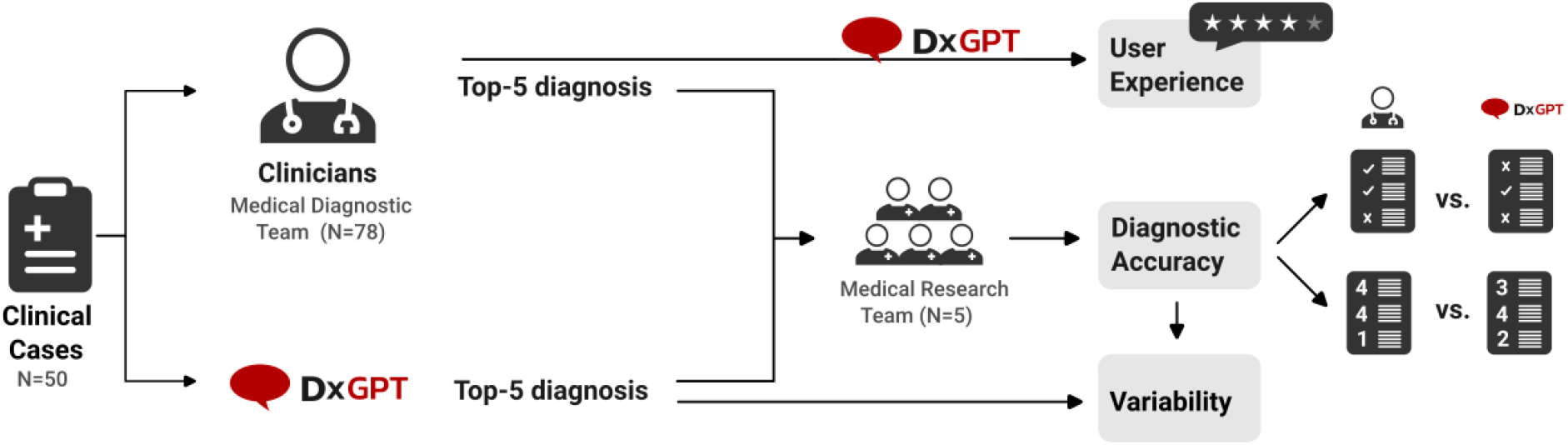
Schematic representation of the study. Real clinical cases reflecting the diversity of a children’s hospital were collected. Clinicians and DxGPT (LLM-based diagnostic tool) provided primary and up to 4 additional diagnoses, which were evaluated blindly by the Medical Research Team. The study also evaluated DxGPT’s performance variability and clinician user experience.

The diagnostic accuracy, measured as the proportion of clinical cases where the correct diagnosis was included among the 5 diagnostic options (Top-5), showed comparable and not statistically significant differences between the two groups. Top-5 diagnostic accuracy was 65% for clinicians and 60% for DxGPT (Figure 2A, eFigure 2 from Supplement 1 and eTable 5 from Supplement 2). We divided clinical cases according to disease frequency and found that common diseases showed higher diagnostic accuracies than rare diseases (two-tailed Fisher’s exact Test: *P* = 7.65×10^−9^; Figure 2B). In both cases, even though clinicians showed slightly higher diagnostic accuracy, this difference was not statistically different from that of DxGPT. Moreover, our results confirm that additional information enhances the accuracy of the diagnostic process, though this is only statistically significant in clinicians (McNemmar’s Test: *P* = 2.25×10^−3^; eFigure 3 from Supplement 1).

**Figure 2.**
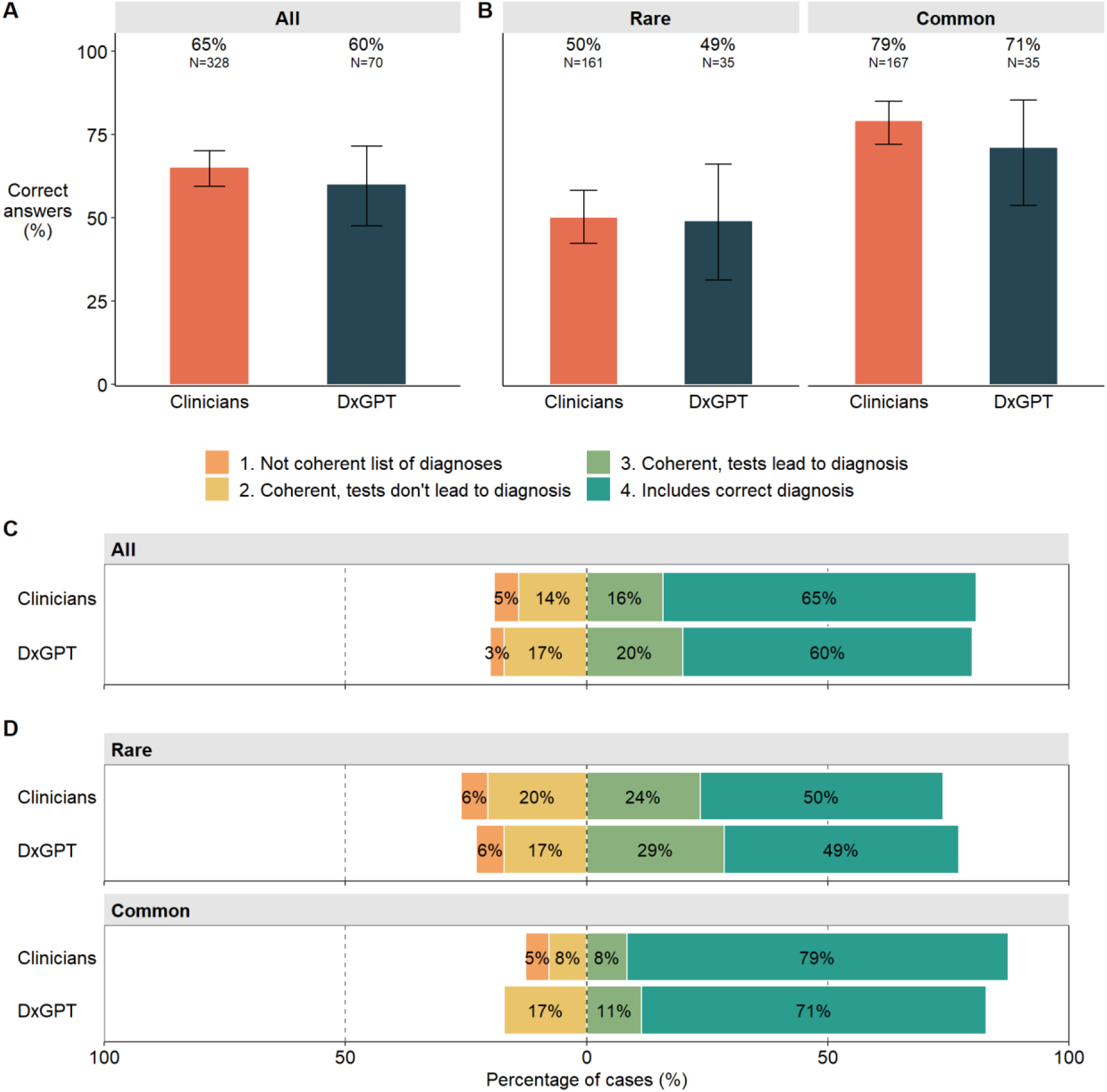
Performance of clinicians vs DxGPT. Comparison of diagnostic accuracy (percentage of correct answers) between diagnostic groups in the dichotomous evaluation (correct diagnosis vs incorrect diagnosis) for (A) all clinical cases, and (B) disease frequency (rare vs common). Comparison of 4-point Likert scale results between diagnostic groups for (C) all clinical cases, and (D) disease frequency.

Remarkably, we also confirmed that the complexity categories the clinical cases had been classified and used for case randomization (see Data extraction and case description) showed different degrees of diagnostic accuracy (eFigure 4 from Supplement 1; two-tailed Fisher’s exact Test for all clinical cases, segregating by diagnostic group: *P <* 1.24×10^−4^). As expected, diagnostic accuracy was reduced as case complexity increased. These tendencies were also comparable between clinicians and DxGPT.

Out of 70 clinical cases, DxGPT and all clinicians correctly diagnosed 40% (N = 28) and misdiagnosed 11% (N = 8). Six cases had opposing results, being correctly diagnosed by one group but not the other. Both DxGPT and clinicians failed on medium or high complexity cases, including six rare and two common conditions. Extended information did not improve accuracy, except in one case of leukodystrophy, where one clinician succeeded with simple information but failed with extended information. DxGPT correctly diagnosed two rare and one common high-complexity case that clinicians missed, while clinicians correctly diagnosed one rare and two common cases of varying complexity that DxGPT failed to diagnose (eTable 6 from Supplement 1).

While diagnostic accuracy assessed in terms of hits and misses provides insight into the performance of DxGPT as a diagnostic tool, it inevitably leaves all missed diagnoses unexamined. To better understand the nuances on undiagnosed cases, we reevaluated all responses using a 4-point Likert scale (Figure 2C-D, eFigures 5 and 6 from Supplement 1 and eTable 5 from Supplement 2). Similarly, to the results obtained with diagnostic accuracy, both diagnostic tools demonstrated parallel trends. Although DxGPT seems to recover more coherent diagnoses leading to the correct one (Likert scale value of 3), these numbers are not significantly different from that found in clinicians (Cochran’s Armitage Test: *P* > 0.05).

### Assessing the variability of DxGPT

We assessed how the variability of the model could affect the responses provided by DxGPT and its diagnostic accuracy. For that, we queried each clinical case in DxGPT three times (eTable 7 from Supplement 2). Our results show that whereas DxGPT is not consistent in its five diagnostic responses and thus shows variability (median pairwise Jaccard = 0.67; Figure 3A), if we focus only on the first diagnosis (Top-1), it is more consistent (median pairwise Jaccard = 1; eFigure 7 from Supplement 1). Remarkably, there is a tendency towards more consistency in the responses to rare diseases though it is not statistically significant (Figure 3B and eFigure 7 from Supplement 1). When we analyzed the impact of response variability of DxGPT on diagnostic accuracy – specifically, whether such variability would result in the loss of the correct diagnostic option– we found that despite some inconsistencies (Figure 3A and 3C), these affect less than 5% of the cases.

**Figure 3.**
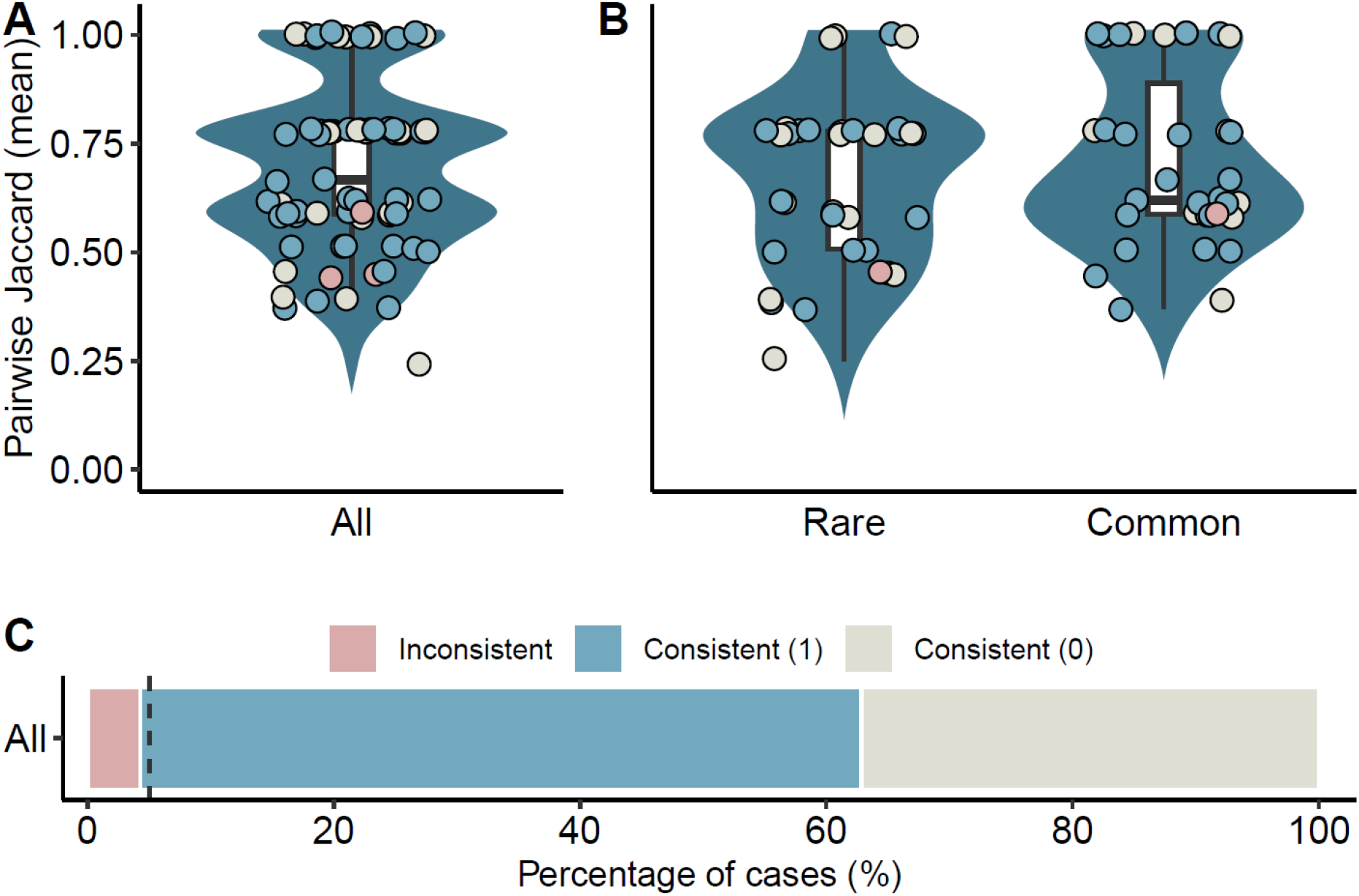
DxGPT variability and its effect on diagnostic accuracy. Distribution of mean pairwise Jaccard coefficient calculated for the Top-5 diagnoses in each clinical case for (A) all clinical cases (n = 70) and (B) segregating by disease frequency (n = 35 clinical cases each). Points were jittered to simplify visualization. Values indicate the tendency to overlap between triplicate diagnoses for each clinical case, ranging from 0 –no overlap-to 1 –complete overlap. The color of each point indicates the effect of DxGPT’s variability on the diagnostic accuracy as in (C). (C) The effect of DxGPT’s variability on the diagnostic accuracy. Percentage of consistent (n = 67) and inconsistent (n = 3) diagnoses. The vertical dashed line indicates 5%. Consistent (1) and (0) denote consistency in the presence or absence of the correct diagnosis, respectively.

### UX evaluation of DxGPT

To evaluate the usability of the diagnostic tool in a clinical setting, clinicians completed a six-question survey after using the DxGPT platform (eTable 8 from Supplement 2). The 48 respondents provided favorable reviews, with a mean overall experience rating of 3.9/5, usefulness for rare or complex cases rated at 4.1/5, and ease of use rated at 4.5/5 (Figure 4A). Higher ratings indicated a more favorable attitude towards DxGPT. No significant differences were found between experience levels, though junior clinicians were generally more optimistic. Clinicians also provided open-ended feedback on their likes, dislikes, and suggestions for improvement summarized in Figure 4B.

**Figure 4.**
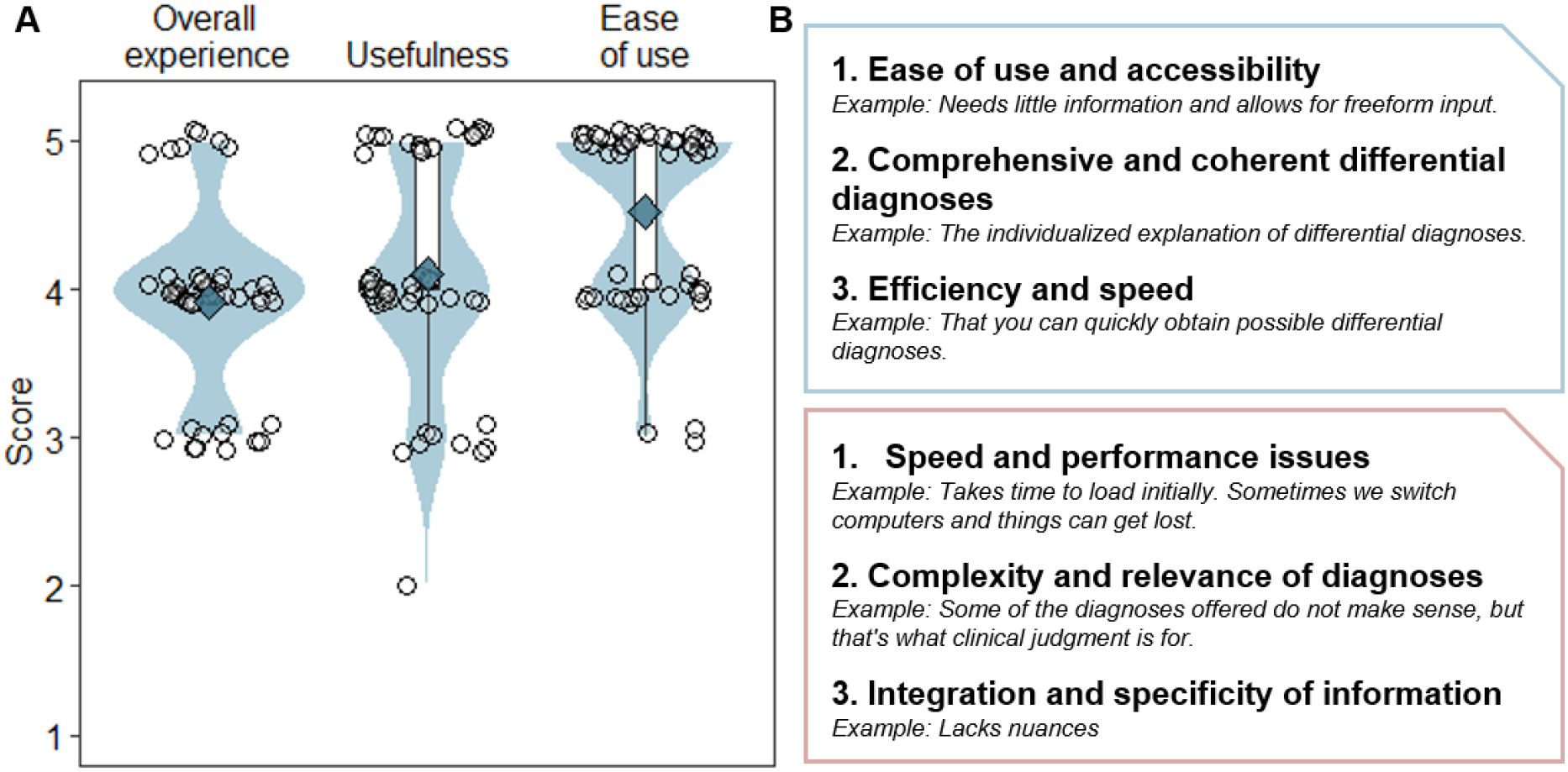
Clinicians’ user experience assessment on DxGPT. (A) Distribution of the scores given for the numerical categories assessed. Empty circles indicate individual responses and blue diamonds indicate the mean score for each category. Points were jittered to simplify visualization. (B) Summary of positive (top) and negative (bottom) feedback regarding DxGPT.

## Discussion

This study provides preliminary evidence on the potential utility of LLMs in aiding differential diagnosis in pediatrics. Our results indicate that the diagnostic accuracy of DxGPT is comparable to that of clinical experts, highlighting its clinical utility and accuracy. Notably, DxGPT’s utility extends beyond common diseases, showing promising results in helping to diagnose rare diseases.

The diagnostic accuracy of DxGPT aligns with other studies comparing LLM-based tools to medical professionals. For instance, a study found GPT-4’s diagnostic accuracy comparable to that of board-certified physicians, correctly diagnosing around 98% of cases in a set of 45 clinical vignettes ^6^. Similarly, other studies on GPT-4 found that it matched experts in ophthalmology and significantly outperformed non-specialist doctors ^7^, it achieved 76.4% in the clinical cases part of the UK Membership of the Royal College of Physicians test ^8^, and obtained a perfect score on the National Board of Medical Examiners (NBME) ^9^.

Most studies use synthetic clinical cases to assess diagnostic accuracy, but they may not capture the complexity of real-world patient encounters. We address this by using clinical cases based on existing pediatric patients, which are inherently more complex and non-stereotypical. Compared to the previously cited studies, ours show lower overall accuracy, which we attribute to the use of real clinical cases. In fact, when we specifically examined low-complexity cases, both clinicians and DxGPT achieved accuracies like those reported in the literature ^6,10^.

In our study, additional information increased the performance of clinical experts and DxGPT with an increase similar to that of Rao et al. ^5^. The differences in DxGPT were not statistically significant due to the low number of cases where this information was available. These results could be improved if semantic reasoning instead of numerical values had been provided as extended information for specific observations like those provided in laboratory results. As LLMs have not yet reached human-level conversational abilities, this limits their ability to fully capture the complexity of medical interactions ^11^.

Our study found that both DxGPT and clinicians had higher diagnostic accuracies for common diseases than rare diseases, reflecting the inherent challenges of diagnosing rare and complex conditions. DxGPT’s ability to correctly diagnose high-complexity cases that were missed by clinicians highlights the potential for LLM-based tools to offer valuable complementary insights in complex diagnostic situations.

LLMs are inherently non-deterministic due to the probabilistic nature of their architecture ^12^. Despite this, we found that DxGPT demonstrated minimal response inconsistencies among replicates, highlighting its robustness. As LLMs continue to evolve and surpass their previous capabilities ^2,10,13,14^, and with the potential for specialized medical training and refinement ^15^, we anticipate future models will further improve diagnostic accuracy, potentially exceeding the performance of clinical experts. However, regardless of the specific performance metrics of any LLM-based tool^16^, correct tool usage remains crucial. Effective prompting strategies ^17,18^ and appropriate application by users are essential for optimizing the performance of these tools ^19,20^.

It is important to consider the inherent limitations in this study. First, the clinical cases, though based on real-world scenarios, required detailed patient information and were limited in number. Moreover, this set of cases reflects the population specific of this hospital. Second, all clinical cases were presented in Spanish, which is not the primary language for most LLMs, including GPT-4. While GPT-4 has demonstrated proficiency in multilingual tasks ^21,22^, we did not evaluate DxGPT’s performance across different languages. Future studies should address these limitations by comparing DxGPT’s performance across multiple languages to ensure its effectiveness in diverse linguistic contexts. Third, we did not evaluate DxGPT’s ability to interpret and weigh clinical risks accurately. Although the Likert score serves as a safety measure, the study’s scope necessitates a comprehensive review covering a broader range of diseases and information levels. Finally, other problems associated with LLMs in medicine include incorrect and potentially unsafe responses, perpetuation of biases in training data, patient data privacy risks ^23–25^, and potential reduction in critical thinking and decision-making skills among medical professionals and students. All of these underscore the need for ongoing research and careful implementation ^8,9,20^.

## Conclusions

To conclude, DxGPT showed similar accuracies to medical doctors from a pediatric reference hospital, indicating its potential supporting clinicians in the initial diagnostic steps for challenging pediatric cases, supported by favorable user experiences. The adaptability and scalability of these AI tools could benefit regions with limited access to medical specialists, improving global healthcare quality and patient outcomes. However, further complementary studies are needed before fully integrating these models into clinical practice.

## Data Availability

Clinical cases with deidentified data without patient's sensitive and confidential information are available upon reasonable request to the corresponding author. Code to replicate analyses and figures and eTables 1, 5, 7 and 8 will be made public upon publication.

https://github.com/maralest/DxGPT_HSJD_Analysis

## Acknowledgements

We would like to thank the clinicians from Hospital Sant Joan de Déu who generously volunteered their time and expertise to participate in this project.

## Author Contributions

*Concept and design*: Launes, Martínez, Isla, Martínez Roldán, Garcia-Cuyas, Esteller-Cucala.

*Acquisition, analysis, or intepretation of data*: Alvarez-Estape, Cano, González Grado, Pino, Aldemira-Liz, Launes, Esteller-Cucala.

*Drafting of the manuscript*: Alvarez-Estape, Launes, Esteller-Cucala.

*Critical review of the manuscript for important intellectual content*: All authors.

*Statistical analysis*: Alvarez-Estape, Esteller-Cucala.

*Administrative, technical, or material support*: Gonzálvez-Ortuño, do Olmo, Logroño,

*Supervision*: Alvarez-Estape, Launes, Esteller-Cucala.

## Data Sharing Statement

Clinical cases with deidentified data without patient’s sensitive and confidential information are available upon reasonable request to the corresponding author. Code to replicate analyses and figures and eTables 1, 5, 7 and 8 (Supplement 2) will be made public upon publication.

## References

1. Molster C, Urwin D, Di Pietro L, et al. Survey of healthcare experiences of Australian adults living with rare diseases. Orphanet J Rare Dis. 2016;11(1). doi:10.1186/s13023-016-0409-z

2. Thirunavukarasu AJ, Ting DSJ, Elangovan K, Gutierrez L, Tan TF, Ting DSW. Large language models in medicine. Nat Med. 2023;29(8):1930–1940. doi:10.1038/s41591-023-02448-8

3. Meng X, Yan X, Zhang K, et al. The application of large language models in medicine: A scoping review. iScience. 2024;27(5):109713. doi:10.1016/j.isci.2024.109713

4. do Olmo J, Logroño J, Mascías C, Martínez M, Isla J. Assessing DxGPT: Diagnosing rare diseases with various large language models. medRxiv. Published online May 9, 2024. doi:10.1101/2024.05.08.24307062

5. Rao A, Kim J, Kamineni M, et al. Evaluating GPT as an adjunct for radiologic decision making: GPT-4 versus GPT-3.5 in a breast imaging pilot. J Am Coll Radiol. 2023;20(10):990–997. doi:10.1016/j.jacr.2023.05.003

6. Ito N, Kadomatsu S, Fujisawa M, et al. The accuracy and potential racial and ethnic biases of GPT-4 in the diagnosis and triage of health conditions: Evaluation study. JMIR Med Educ. 2023;9(1):e47532. doi:10.2196/47532

7. Thirunavukarasu AJ, Mahmood S, Malem A, et al. Large language models approach expert-level clinical knowledge and reasoning in ophthalmology: A head-to-head cross-sectional study. PLOS Digit Health. 2024;3(4):e0000341. doi:10.1371/journal.pdig.0000341

8. Maitland A, Fowkes R, Maitland S. Can ChatGPT pass the MRCP (UK) written examinations? Analysis of performance and errors using a clinical decision-reasoning framework. BMJ Open. 2024;14(3):e080558. doi:10.1136/bmjopen-2023-080558

9. Abbas A, Rehman MS, Rehman SS. Comparing the performance of popular large language models on the national board of medical examiners sample questions. Cureus. Published online March 11, 2024. doi:10.7759/cureus.55991

10. Sandmann S, Riepenhausen S, Plagwitz L, Varghese J. Systematic analysis of ChatGPT, Google search and Llama 2 for clinical decision support tasks. Nat Commun. 2024;15(1):1–8. doi:10.1038/s41467-024-46411-8

11. Lucas HC, Upperman JS, Robinson JR. A systematic review of large language models and their implications in medical education. Med Educ. Published online April 19, 2024. doi:10.1111/medu.15402

12. Peeperkorn M, Kouwenhoven T, Brown D, Jordanous A. Is temperature the creativity parameter of large language models? arXiv [csCL]. Published online May 1, 2024. Accessed July 16, 2024. http://arxiv.org/abs/2405.00492

13. Hirosawa T, Harada Y, Yokose M, Sakamoto T, Kawamura R, Shimizu T. Diagnostic accuracy of differential-diagnosis lists generated by generative pretrained transformer 3 chatbot for clinical vignettes with common chief complaints: A pilot study. Int J Environ Res Public Health. 2023;20(4):3378. doi:10.3390/ijerph20043378

14. Hirosawa T, Harada Y, Tokumasu K, Ito T, Suzuki T, Shimizu T. Evaluating ChatGPT-4’s diagnostic accuracy: Impact of visual data integration. JMIR Med Inform. 2024;12:e55627. doi:10.2196/55627

15. Guo E, Gupta M, Sinha S, et al. neuroGPT-X: toward a clinic-ready large language model. J Neurosurg. 2024;140(4):1041–1053. doi:10.3171/2023.7.jns23573

16. Cabitza F, Campagner A, Del Zotti F, Ravizza A, Sternini F. All you need is higher accuracy? On the quest for minimum acceptable accuracy for Medical Artificial Intelligence. In: Macedo M, ed. Proceedings of the 12th International Conference on e-Health (EH2020); 2020. doi:10.33965/eh2020_202009l020

17. Meskó B. Prompt engineering as an important emerging skill for medical professionals: Tutorial. J Med Internet Res. 2023;25:e50638. doi:10.2196/50638

18. Abdullahi T, Singh R, Eickhoff C. Learning to make rare and complex diagnoses with generative AI assistance: Qualitative study of popular large language models. JMIR Med Educ. 2024;10:e51391. doi:10.2196/51391

19. Goh E, Gallo R, Hom J, et al. Influence of a large language model on diagnostic reasoning: A randomized clinical vignette study. bioRxiv. Published online March 14, 2024:2024.03.12.24303785. doi:10.1101/2024.03.12.24303785

20. Rao A, Pang M, Kim J, et al. Assessing the utility of ChatGPT throughout the entire clinical workflow: Development and usability study. J Med Internet Res. 2023;25(1):e48659. doi:10.2196/48659

21. Zhao Y, Zhang W, Chen G, Kawaguchi K, Bing L. How do large language models handle multilingualism? arXiv [csCL]. Published online February 28, 2024. Accessed July 16, 2024. http://arxiv.org/abs/2402.18815

22. Plaza I, Melero N, del Pozo C, et al. Spanish and LLM benchmarks: Is MMLU lost in translation? arXiv [csCL]. Published online May 28, 2024. Accessed July 16, 2024. http://arxiv.org/abs/2406.17789

23. Li H, Moon JT, Purkayastha S, Celi LA, Trivedi H, Gichoya JW. Ethics of large language models in medicine and medical research. Lancet Digit Health. 2023;5(6):e333–e335. doi:10.1016/s2589-7500(23)00083-3

24. Ong JCL, Chang SYH, William W, et al. Ethical and regulatory challenges of large language models in medicine. Lancet Digit Health. 2024;6(6):e428–e432. doi:10.1016/s2589-7500(24)00061-x

25. Deng J, Zubair A, Park YJ, Affan E, Zuo QK. The use of large language models in medicine: proceeding with caution. Curr Med Res Opin. 2024;40(2):151–153. doi:10.1080/03007995.2023.2295411

