## SupplementaryMaterial for "Evaluation of the Clinical Utility of DxGPT, a GPT-4 Based Large Language Model, through an Analysis of Diagnostic Accuracy and User Experience"

### Supplement 1

**eMethods.** Survey on the Use and Evaluation of the DxGPT Tool.

**eFigure 1.** Participant recruitment and case assessment procedures.

**eFigure 2.** Paired evaluation of the performance of clinicians vs DxGPT.

**eFigure 3.** Diagnostic accuracy comparison cases with extended information (N=20).

**eFigure 4.** Evaluation of the performance of clinicians vs. DxGPT per complexity.

**eFigure 5.** Evaluation of Likert scale results of clinicians vs DxGPT.

**eFigure 6.** Evaluation of Likert scale results of clinicians vs DxGPT for cases with extended information (N=20).

**eFigure 7.** DxGPT variability and its effect on diagnosis for the first diagnosis (Top-1).

**eTable 2.** Number of clinical cases by specialty considered for the study. Number proportional to their occurrence at the hospital.

**eTable 3.** Number and proportion of genders for each disease frequency (rare and common).

**eTable 4.** Number and proportion of cases per complexity and disease frequency (rare and common).

**eTable 6.** Clinical Cases that were misdiagnosed by both groups (DxGPT and clinicians) and correctly diagnosed by one of the groups and not the other.

### eMethods

**Survey on the Use and Evaluation of the DxGPT Tool.** All questions were originally asked and answered in Spanish.

1. ¿Cómo calificarías tu experiencia general con DxGPT? (1 Muy mala, 5 Muy buena)
   *How would you rate your overall experience with DxGPT? (1 Very bad, 5 Very good)*
2. ¿Qué tan útil encuentras el uso de DxGPT4 en tu práctica diaria, especialmente para casos raros o difíciles? (1 Nada útil, 5 Muy útil)
   *How useful do you find the use of DxGPT4 in your daily practice, especially for rare or difficult cases? (1 Not useful, 5 Very useful)*
3. ¿Qué tan fácil te ha parecido usar DxGPT? (1 Muy difícil, 5 Muy fácil)
   *How easy have you found it to use DxGPT? (1 Very difficult, 5 Very easy)*
4. ¿Qué te ha gustado más de DxGPT? (Respuesta abierta)
   *What have you liked most about DxGPT? (Open-ended question)*
5. ¿Qué te ha gustado menos de DxGPT? (Respuesta abierta)
   *What have you liked least about DxGPT? Open-ended question)*
6. ¿Hay alguna característica específica o mejora que te gustaría ver en la herramienta? (Respuesta abierta)
   *Is there any specific feature or improvement you would like to see in the tool? Open-ended question)*

#### eFigures

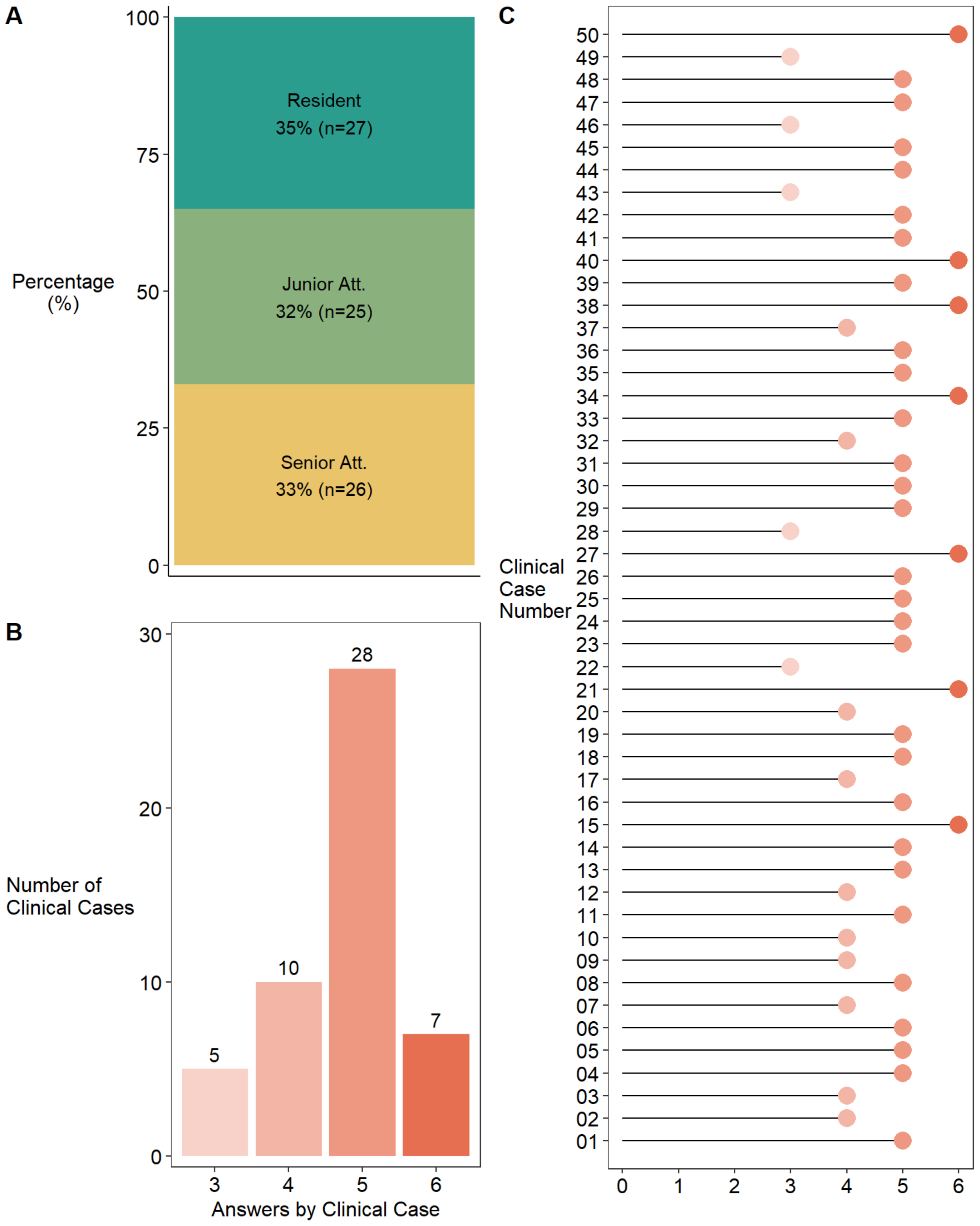


**eFigure 1. Participant recruitment and case assessment procedures.** (A) Percentage of respondents by professional experience (Resident, Junior Attending or Senior Attending). Residents are defined as clinicians in training with 1 to 4 years of experience in clinical practice, Junior Attendings have less than 7 years of experience, and Senior Attendings have 7 or more years of experience in clinical practice. (B) Distribution of the number of answers per clinical case. (C) Number of answers by clinical case.

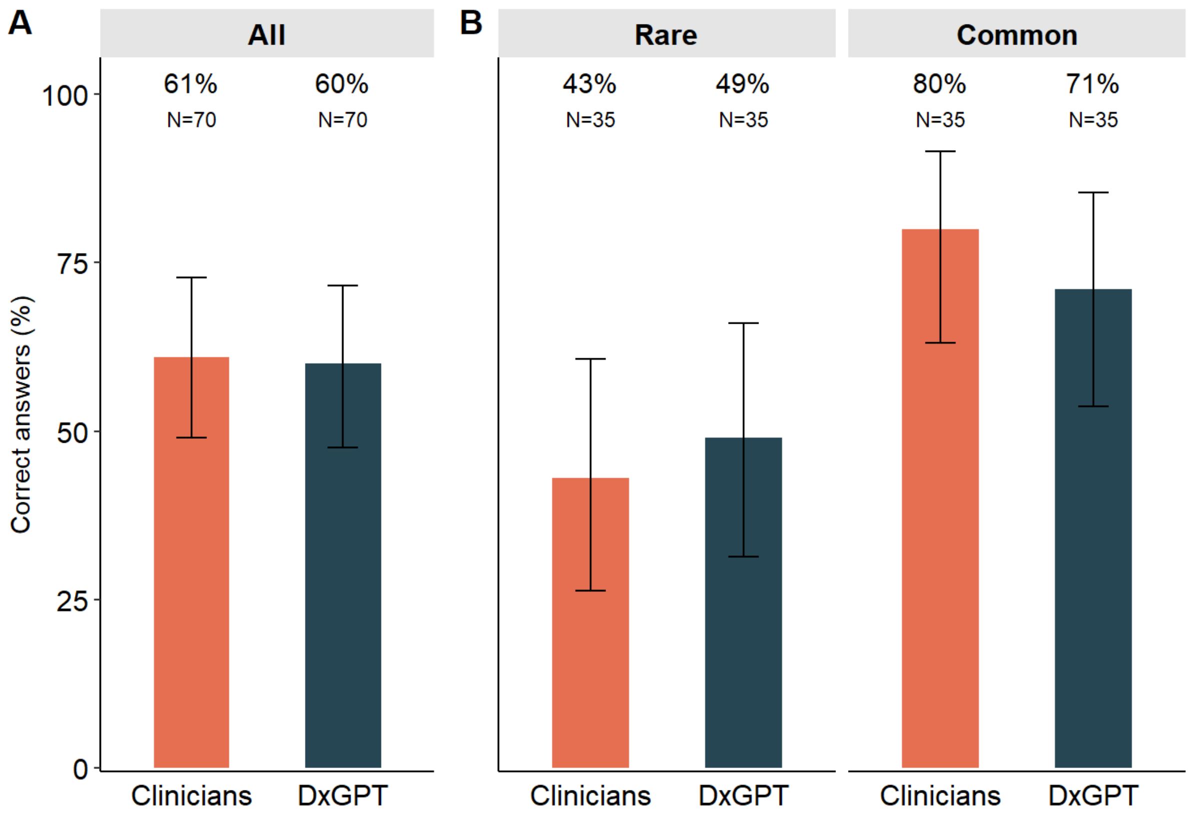


**eFigure 2. Paired evaluation of the performance of clinicians vs DxGPT.** Comparison of diagnostic accuracy (percentage of correct answers) between diagnostic groups in the dichotomous paired evaluation (using the mode of the scores for each clinical case evaluated by clinicians) for (A) all clinical cases, and (B) disease frequency (rare vs common).

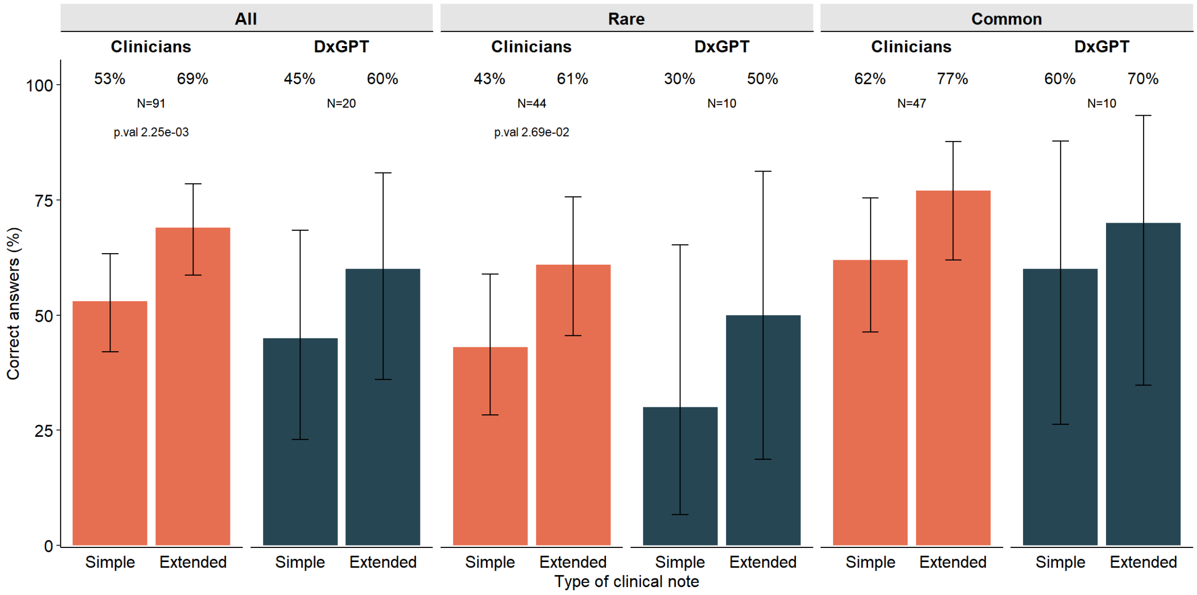


**eFigure 3. Diagnostic accuracy comparison cases with extended information (N=20).** Paired comparison of diagnostic accuracy (percentage of correct answers) for the 20 clinical cases that had extended information for all, rare and common cases.

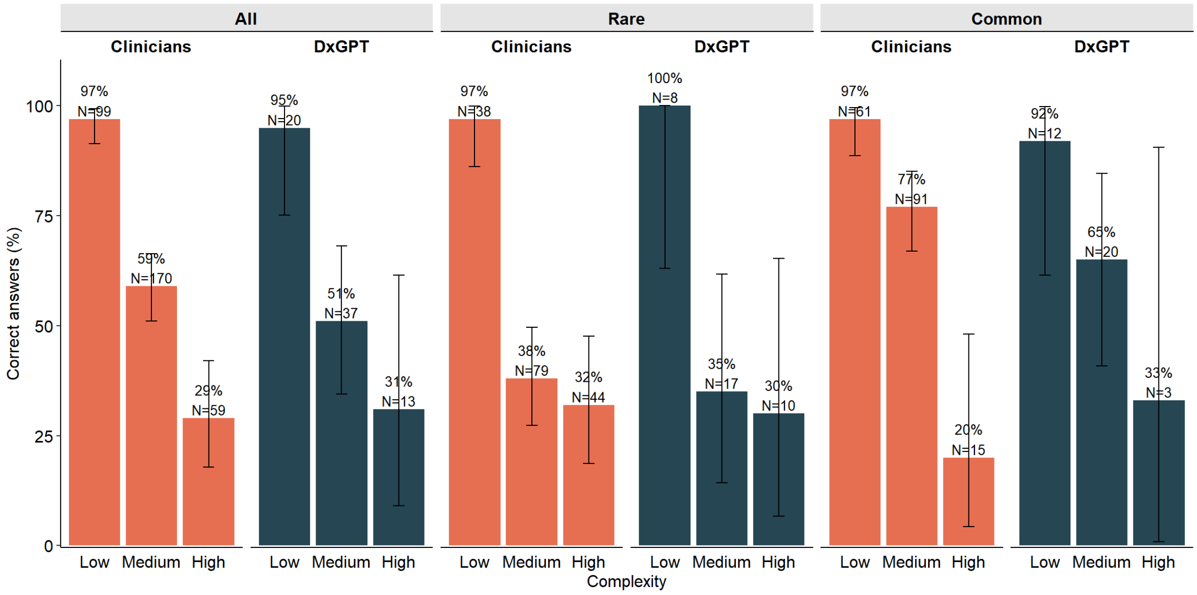


**eFigure 4. Evaluation of the performance of clinicians vs. DxGPT per complexity.** Comparison of diagnostic accuracy (percentage of correct answers) between diagnostic groups in the dichotomous evaluation per complexity level for (A) all clinical cases, and (B) disease frequency (rare vs common).

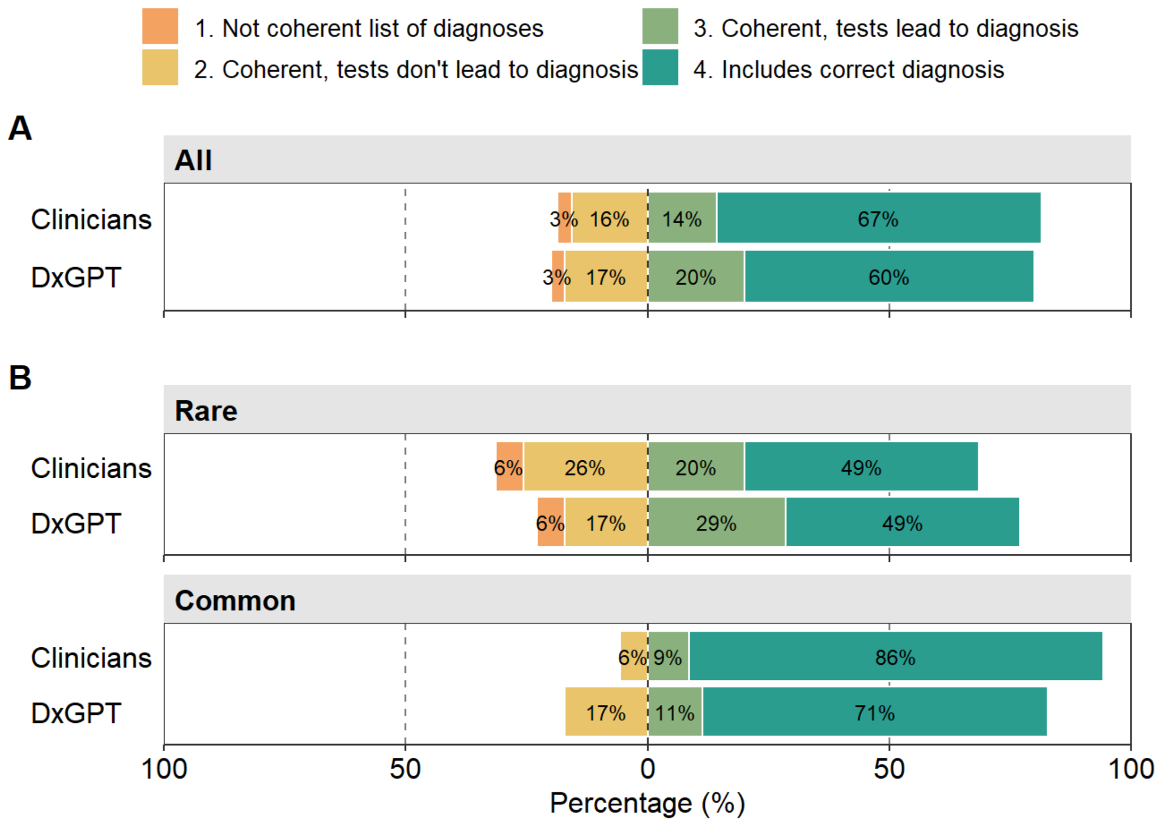


**eFigure 5. Evaluation of Likert scale results of clinicians vs DxGPT.** Comparison of 4-point Likert scale results between diagnostic groups per for (A) all clinical cases, and (B) disease frequency (rare vs common). To ensure paired evaluation, the mode of the Likert scores for each clinical case evaluated by clinicians were used.

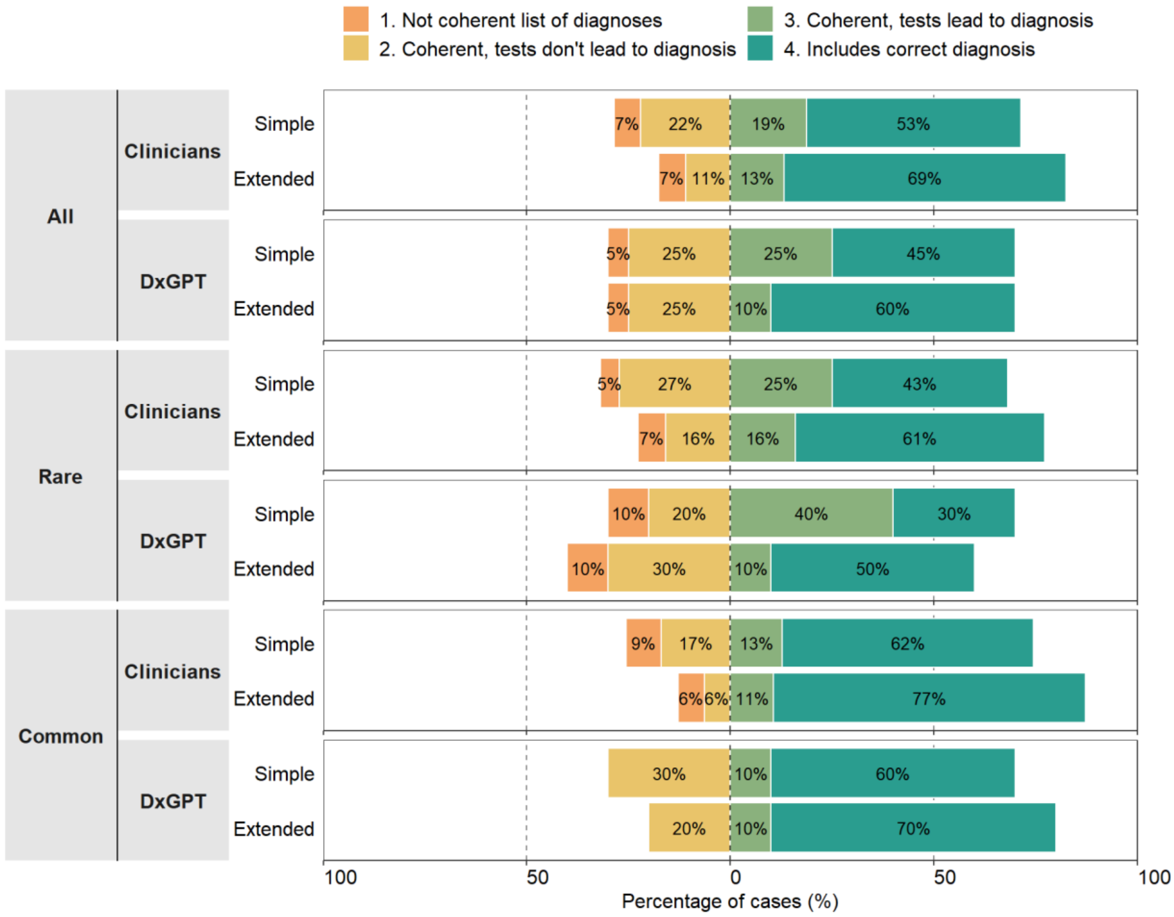


**eFigure 6. Evaluation of Likert scale results of clinicians vs DxGPT for cases with extended information (N=20).** Comparison of 4-point Likert scale results between diagnostic groups per for those cases with extended information. (A) all clinical cases, and (B) disease frequency (rare vs common).

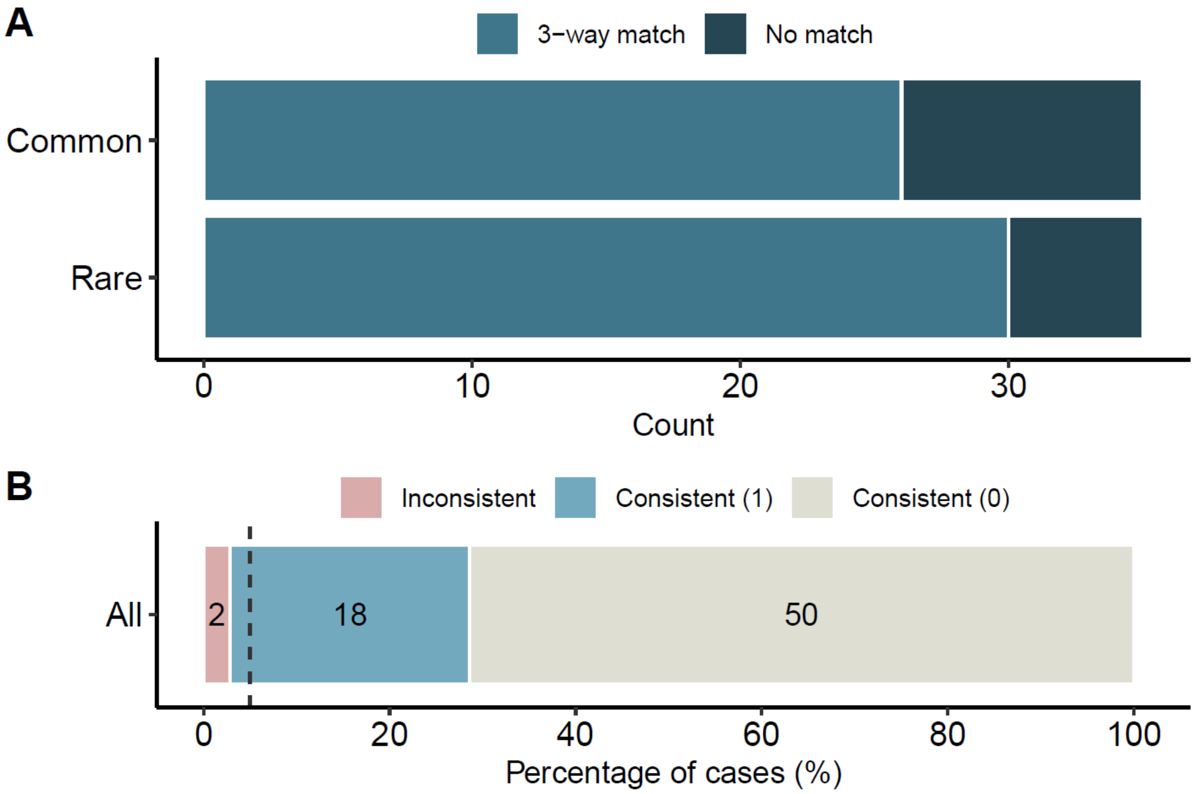


**eFigure 7. DxGPT variability and its effect on diagnosis for the first diagnosis (Top-1).** (A) Counts of Top-1 variability possibilities per disease frequency, where *3-way match* corresponds to a mean Jaccard coefficient of 1, otherwise it is labeled as *No match*. (B) The effect of DxGPT's variability on the diagnostic accuracy for all clinical cases considering the first diagnosis. Percentage of consistent (n = 68) and inconsistent (n = 2) diagnoses. The vertical dashed line indicates 5%. Consistent (1) and (0) denote consistency in the presence or absence of the correct diagnosis, respectively.

#### eTables

**eTable1. Clinical cases with diagnosis, frequency type, complexity, gender and indication whether they have extended information (N = 50).** *See Supplement 2 (xlsx)*

**eTable 2. Number of clinical cases by specialty considered for the study. Number proportional to their occurrence at the hospital.**

| Clinical specialty | Distribution of cases (%) | |
|---|---|---|
| | Theoretical | Real (Simple + Extended) |
| Gastroenterological cases | 20 | 20 |
| Neurological/metabolic conditions | 20 | 19 |
| Oncological cases | 20 | 23 |
| Infectious diseases | 10 | 7 |
| Nephrological conditions | 10 | 10 |
| Rheumatological cases & Immunological conditions | 10 | 9 |
| Others (Endocrinology, Cardiology or Dermatology, among others) | 10 | 13 |
| *Total* | *100* | *100* |

**eTable 3. Number and proportion of genders for each disease frequency (rare and common).**

| Disease frequency | Counts | | Percentage (%) | |
|---|---|---|---|---|
| | Female | Male | Female | Male |
| Rare | 12 | 13 | 48 | 52 |
| Common | 12 | 13 | 48 | 52 |
| *Total* | *24* | *26* | *48* | *52* |

**eTable 4. Number and proportion of cases per complexity and disease frequency (rare and common).**

| Disease frequency | Counts | | | Percentage (%) | | |
|---|---|---|---|---|---|---|
| | Low | Medium | High | Low | Medium | High |
| Rare | 6 | 11 | 8 | 24 | 44 | 32 |
| Common | 10 | 13 | 2 | 40 | 52 | 8 |
| *Total* | *16* | *24* | *10* | *32* | *48* | *20* |

**eTable 5. Top-5 and Likert scale results for clinicians and DxGPT.** *See Supplement 2 (xlsx)*

**eTable 6. Clinical Cases that were misdiagnosed by both groups (DxGPT and clinicians) and correctly diagnosed by one of the groups and not the other.**

| Clinical Case | Simple or Extended | Rare or Common | True Diagnosis | Complexity | Comparison |
|---|---|---|---|---|---|
| HC_04 | Simple | Rare | Congenital diaphragmatic hernia | Medium | All failed |
| HC_08 | Simple | Rare | Williams syndrome | High | All failed |
| HC_09 | Extended | Rare | Aicardi-Goutières syndrome | High | All failed |
| HC_19 | Simple | Rare | Pseudohypoaldosteronism type 1 | Medium | All failed |
| HC_20 | Simple | Rare | Posterior urethral valve | Medium | All failed |
| HC_35 | Simple | Common | Methylmalonic acidemia | High | All failed |
| HC_38 | Simple | Rare | Brain tumor/Diffuse Intrinsic Pontine Glioma | Medium | All failed |
| HC_48 | Simple | Common | Congenital adrenal hyperplasia | Medium | All failed |
| HC_01 | Simple | Rare | Bardet-Biedl syndrome | High | Only DxGPT passed |
| HC_23 | Simple | Rare | Pediatric-onset Graves disease | High | Only DxGPT passed |
| HC_35 | Extended | Common | Methylmalonic acidemia | High | Only DxGPT passed |
| HC_22 | Simple | Rare | Macrophage activation syndrome | High | Only clinicians passed |
| HC_30 | Simple | Common | Cow's milk protein allergy | Low | Only clinicians passed |
| HC_43 | Simple | Common | Acute post-infectious glomerulonephritis | Medium | Only clinicians passed |

**eTable 7. List of diagnosis given by DxGPT in triplicates to assess variability with Top-5 and Top-1 evaluations.** *See Supplement 2 (xlsx)*

**eTable 8. User experience ratings to three 5-point scale questions.** *See Supplement 2 (xlsx)*
